# The undue influence of genetic information on medical students’ treatment decisions

**DOI:** 10.1101/2022.10.31.22281782

**Authors:** Andrew S. Lane, Kate E. Lynch, Mark Arnold, Ilan Dar-Nimrod, James Morandini, Stefan A. Gawronski, Paul E. Griffiths

## Abstract

**Introduction:** Knowledge of the genetic basis of health conditions can influence how the public perceives their own and others’ health. When there are known genetic associations for such conditions, genetic essentialist biases facilitate deterministic thinking and an over-emphasis of genetic causality. This study investigates the role that genetic essentialist biases play in medical decision making.

**Methods:** Third- and fourth- year medical students (N = 102) read a scenario in which a patient presents with gastroenterological symptoms. Half of the students were told that the patient tested positive for HLADQ2 – a gene implicated in, but not deterministic of, coeliac disease. The other half received no genetic information. Students were assessed on their recommendations for investigation and management using a multiple-choice questionnaire. Twenty-two of these students participated in a qualitative follow-up which used semi-structured interviews to explore the reasoning behind students’ responses.

**Results:** Management recommendations differed between the two groups, with those receiving genetic information more likely to recommend a gluten free diet. Recommendations for further investigation did not differ significantly between groups. Interviews suggested that these findings arose despite the students’ good understanding of the common non-deterministic nature of genes, such as HLADQ2. Differences in management recommendations suggest that the inclusion of genetic information unduly biased students towards a premature diagnosis of a serious health condition–coeliac disease. Follow-up interviews introduce the possibility that observed manipulation-based differences may have been based on anticipated expectations of examiners. Thus, research in clinical settings is needed to ascertain whether genetic essentialist biases are implicitly influencing medical students under examination conditions or whether they react to the assumptive genetic biases of examiners.

**Conclusion:** The present findings indicate that genetic essentialist biases may affect clinical decision making of senior medical students. While the findings may also arise as an artifact of a conscious exam-taking strategy, there are substantial arguments against this interpretation.

## Introduction

Diagnostic errors are common across all medical disciplines, resulting in potential and actual patient harms, increased health system costs, and reduction of patient trust (Graber et al., 2005). Cognitive biases can significantly affect diagnostic reasoning and and can result in diagnostic error (Bornstein & Emler, 2001; Gopal et al., 2021). Such biases in clinical practice can occur as a result of the manner in which information is presented (O’Sullivan & Schofield, 2018), systemic prejudices directed towards a social or cultural group (e.g. Chiaramonte & Friend, 2006), or the practitioner’s mental state (Loewenstein, 2005). At the time of writing the Centre of Evidence Based Medicine has indexed over 60 different kinds of biases in their ‘Catalogue of Bias’ (*Catalogue of Bias*, 2017). Many of these have not yet been explored in a clinical context.

Recent work in lay populations illustrates the effect that genetic information has on people’s beliefs, decisions, affect, and behaviours. These effects can be explicated through the Genetic Essentialist Framework (GEF). Myriad of phenomena (e.g., traits, health conditions) perceived to have genetic associations are viewed as more predetermined. When both genetic and environmental causes can be identified, individuals often tend to prioritise the causal role of genes, and perceive those traits as more immutable and inevitable (Dar-Nimrod & Heine, 2011; Heine et al., 2017). These biases can influence the perception of self and others (Dietrich et al., 2006; Kvaale et al., 2013; Lebowitz & Ahn, 2014), behaviour (Dar-Nimrod et al., 2014; Ahn & Lebowitz, 2018; Lebowitz et al., 2021), and cognition (Dar-Nimrod & Heine, 2006). For instance, individuals who received feedback about their own genetic predispositions to depression reported reduced feelings of control (Birchwood et al., 1993), less confidence in their ability to cope (Lebowitz & Ahn, 2018), and remembered more depressive symptoms in their recent past (Lebowitz & Ahn, 2017; Ahn et al., 2020).

Genetic information influences the perceived efficacy of medical treatments from the patient perspective. When medical conditions are thought to have a genetic component, non-biogenic treatments, such as psychotherapy, diet or exercise are often discounted (Iselin & Addis, 2003; Ahn & Lebowitz, 2018). This, in combination with the fact that biased decision making can also be elicited via diagnostic suggestion from patients (Durning et al., 2012), suggests that genetic essentialist biases could impact clinical decision making in a world in which patients have increased access to their personal genetic information.

To understand how genetic essentialist biases might play out in a clinical setting, this study examined the influence of genetic information on the clinical decision-making of final-year medical students. The medical students were given genetic information in the form of medical vignettes around a patient presenting with gastrological symptoms. These symptoms could have been caused by coeliac disease, but were likely to be due to more common diseases.

HLADQ2 is a necessary but not sufficient criterion for the development of coeliac disease. Whereas 30-40% of European-descent persons carry the HLADQ2 (or HLADQ8) allele, only around 1% have coeliac disease (Husby et al., 2012). To better understand how genetic information influences students’ clinical decision making, we further explored their educational experience using semi-structured interviews to understand the reasoning behind students’ responses. This included how expectations and experiences of medical school and life as a practitioner influenced their responses in the initial experiment.

## Methodology and methods

### Participants and recruitment

In phase 1, 482 senior medical students from the Sydney Medical Program were offered the opportunity to take part in a 60-minute revision session on gastroenterology. The students were two weeks from undergoing their summative barrier examinations, ensuring that their knowledge levels were optimal prior. The final sample of 102 students (60 male, 52 female; postgraduate medical students, consisting of 17 third-year and 85 fourth-year students in the University of Sydney Medical Program) were randomly allocated to read one of two hypothetical cases (henceforth the scenarios). Participants ranged in age from 23 to 43 (*M =* 26.39 years; *SD* = 3.09).

Under examination conditions in the form of an online learning module, participants read one of two almost identical realistic scenarios describing a patient with a 3-month history of lethargy, occasional diarrhoea, and confabulated feelings. The sole difference between the scenarios was that a HLADQ2 positive result from an at-home genetic test was included in one scenario. Following this, participants completed ten multiple-choice questions, including two specific questions regarding 1) what the appropriate follow-up investigation would be; and 2) what the appropriate management would be (appendix 1, questions 7 and 9 respectively). Following the completion of the quiz, participants were fully debriefed about the the module being a part of a research endeavour to investigate the effect of added genetic information.

Phase 2 facilitated a qualitative phenomenological exploration of participants’ educational experience, focussing on their their responses to the relevant multiple-choice questions, using a mixture of face-to-face focus-groups or personal interviews (based on students’ preference and availability). The sample size (N = 22) was derived using thematic saturation considerations (Moser & Korstjens, 2018).

Each focus-group or interview was conducted 2-3 weeks after the initial study, giving participants the opportunity to reflect on their experiences. Semi-structured interviews were conducted, audio recorded, and transcribed. Data was analysed using Interpretative Phenomenological Analysis (IPA) (Smith et al., 2009), a data analysis method designed to assess, among other, how educational sense-making translates into clinical practice (Lane & Roberts, 2020). IPA aims to understand the participant’s lived experience during the educational module to provide insight that may not be captured using alternative research methods. In IPA studies, small homogenous samples of participants are used and systematically analysed to identify patterns of convergence and divergence (Smith et al., 2009). The results are presented in the form of a narrative account where the researcher’s interpretation is supported by quotations. The analytical process in IPA is an iterative and inductive cycle that starts by examining the particular and being descriptive and then ascends towards examining the shared and being interpretive. The following is a brief description of the process: Close line by line reading of transcripts, becoming familiar with their content, data reduction through descriptive and conceptual coding of the text, identification of emergent patterns and relationships between codes in light of the research question, code reduction and recombination into emergent sub-themes, development of a structure illustrating the relationships between themes.

Ethics application was approved by the University of Sydney HREC, protocol number 2019/239

## Results

### Phase 1

To explore whether providing information about HLADQ2 influenced treatment decisions, a chi square test assessed the ratio of the students who (erroneously) opted to recommend “gluten free diet” as the most appropriate management of the patient symptoms, as a function of the inclusion of genetic information (indicating that they have likely prematurely diagnosed the patient with coeliac disease). The data revealed that 62.0% of the students who were not told about the HLADQ2 opted to recommend the diet, whereas 84.6% of the students provided with the HLADQ2 information did, χ^2^(1) = 6.70, p = .01. The students provided with the HLADQ2 information (63%) were also more likely to erroneously recommend coeliac-related treatments than those who were not (50%), but this difference did not reach significance, χ^2^(1) = 1.88, p = .17.

To ascertain whether the differences observed above were a function of overall better understanding of the medical case among the group of students who were randomly assigned to *not* receive the HLADQ2 information, a *t*-test was performed on the aggregated scores of the eight questions that should not have been affected by that information. The mean number of correct answers among the students who saw the HLADQ2 information (*M = 3*.*77, SE* = 0.21) did not significantly differ from those who did not see this information (*M = 3*.*54, SE* = 0.23), *t*(100) = 0.75, p = .46.

### Phase 2

In phase 2, 22 students were divided into four focus-groups of four students each, and six individual interviews. Analysis of the data demonstrated two superordinate themes: ‘Preparation for future practice’ and ‘My genetic understanding’. Each superordinate theme was made up of two separate themes. The superordinate themes and themes are shown in Figure 1.

**Figure 1.**
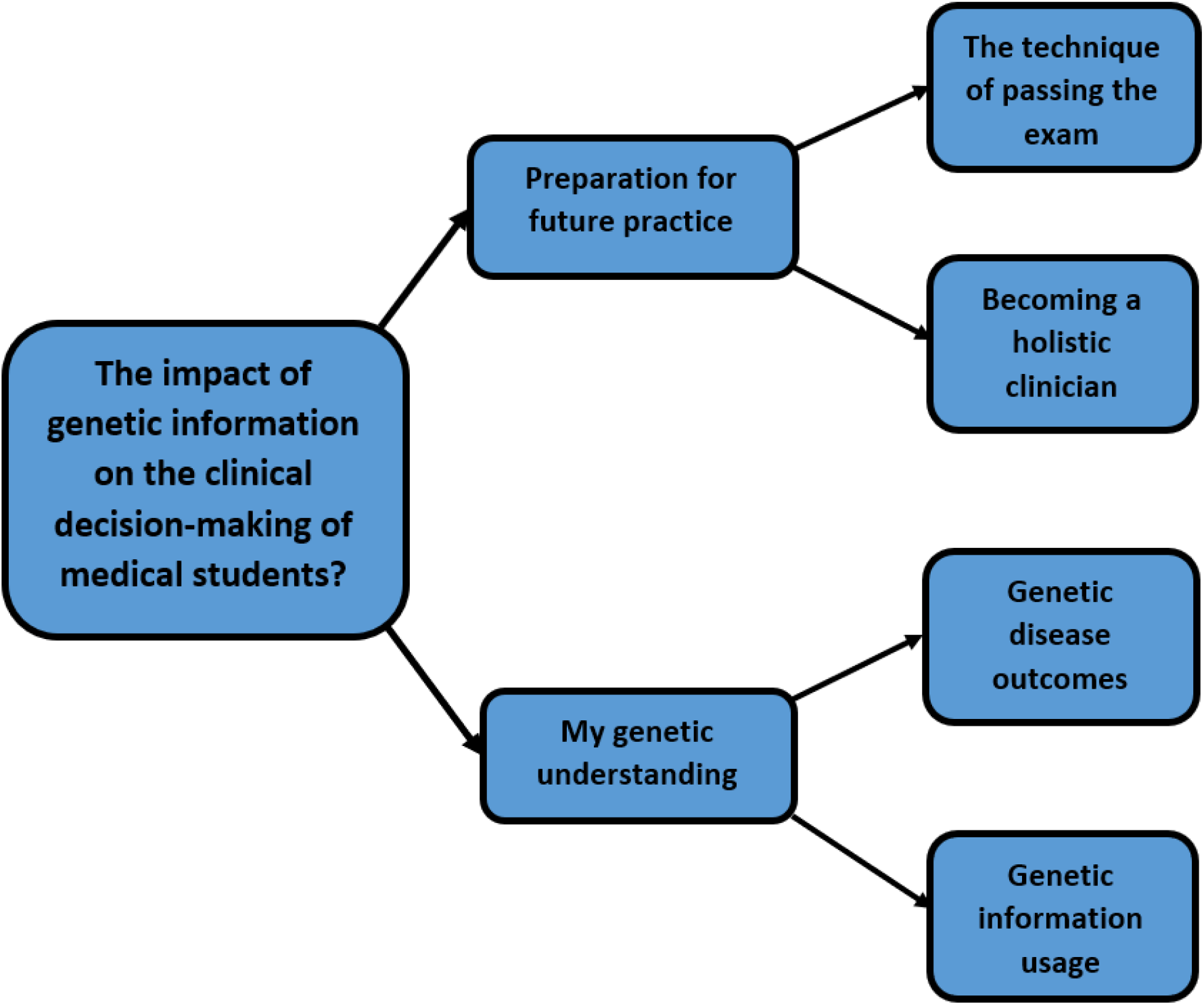
Superordinate themes and themes.

### Superordinate theme 1: Preparation for future practice

This superordinate themes represents the students’ experiences of their medical training, which seem to have taught them to behave differently in the classroom and in the clinical practice. It was made up of two separate themes: 1) the technique of passing the exam, which described the students aiming to identify examiners’ expectation rather than offer a more appropriate clinical recommendation, and 2) becoming a holistic clinician, which described the students’ recognition of their apprenticeship career stage. Boxes 1 and 2 show a selection of supportive quotes for the two themes described above.

#### Box 1

Supportive quotes for theme 1 ‘the technique of passing the exam’

*‘Yeah. No. I’m just trying to phrase it in a way that makes sense. I think the way multiple choice questions are set up, you have to utilise all the information that’s given to you. And if you’re given a piece of information, you can’t disregard it as not important. So with all of those and considering them as important factors, then I would be leaning towards the IGA*.*’*

*‘Well, I mean personally for me I have like two different modes. So I have an exam mode, like an assessment mode, and a clinical mode. So if I were taking it as a – like an assessment, I would see the genetic information and I would think is the question trying to get me to think about a certain diagnosis’*

*‘I also had the genetic and I think I did put down gluten-free diet, but I think it was because of the fact that these were multiple choice questions. In this context I felt like – because it was mentioned, that that’s what I needed – that I had to understand that this was coeliac disease*.*’*

*‘Yeah. And I think it’s something that most medical students are aware of is that there is a big distinction between how you would do something on the wards and how you would do it in an exam, and giving people different stems’*

#### Box 2

Supportive quotes for theme 2 ‘becoming a holistic clinician’

*‘I think any written exam is always going to have the same issue where people are only given a very finite amount of information, and they’re not allowed to seek clarification on it, and to respond to the information they’re given. So I’m a big supporter of on the ward, more clinical-style assessment*.*’*

*‘Partly in principle and partly because I know that if I give them what they want, and then it turns out that they wanted the on the wards question, like the response, then I would be furious with myself. But I think I drop a good five per cent every exam I sit by answering things on principle*.*’*

*‘I think in the immediate context, as an intern, I would probably go and do a first aid of rehydration and recovery because that is what we are taught to do acutely, and then go about with investigations, history exam, and consulting a senior clinician’*

*‘I think it’s important because then you get that kind of instinct, the clinical instinct, but you also have like the locations and the clinical information which help us to refine the clinical instinct. Yeah, so by – I know that when we’re being assessed the assessment purely wants us to make certain connections, and so I think okay when I see this clue, what kind of change should I make and what kind of like relationship do they want me to form?’*

### Superordinate theme 2: My genetic understanding

This superordinate theme represents the students’ understanding of the complexity of genetic aetiology and its associated role in health and disease. It was made up of two separate themes 1) genetic disease outcomes, which described the students’ recognition of the influence of genetic markers on disease, and 2) genetic information usage, which described the students’ hesitancy to use genetic information as a diagnostic tool. Boxes 3 and 4 show a selection of supportive quotes for the themes.

#### Box 3

Supportive quotes for theme 1 ‘genetic disease outcomes’

*‘I’ve had very good experiences with rheumatologists where it’s useful for excluding things, certainly, but positive results are not always indicative of guaranteed disease. So I think I’ve seen that – had that red herring there a few too many times to just go, oh, yep, they’ve got these hundred per cent*.*’*

*‘Because common things are common. So even if – even if someone has had a rare genetically - a gene related illness it doesn’t necessarily – even if the story fits, it doesn’t necessarily mean that they have that or are more likely to have it, it gives more of an indication*.*’*

*‘I think we shouldn’t make them afraid of genetic information. I think it should be told that it’s not absolute. Some people have different types of HLA genes and they’re predisposed to different – there’s a correlation. The correlation versus causation type, I think that needs to be stated*.*’*

#### Box 4

Supportive quotes for theme 2 ‘genetic information usage’

*‘We also don’t tend to do a huge amount of rheumatology, which is where genetics and those tests tend to count the most. So I know for me I tend to rely on like clinical judgment more than genetic information*.*’*

*‘I would look through all that first and then if that didn’t fit, then I probably wouldn’t rely on the genetic test, or the genetic test would not be as accurate, whereas if it fit and the genetic test came back as positive, then for me it’s more confirmation, as opposed to directly guiding me into it and also I often because in the medical program we don’t tend to do a huge amount of genetics*.*’*

*‘So for me genetics is more confirmatory, and that’s only because I’m not familiar with it. I think if I had a better understanding of genetics, I would be able to kind of use of it, whereas for me because it’s uncomfortable, and unfamiliar, it’s only really used to confirm*

## Discussion

As predicted, medical students exposed to genetic information were significantly more likely to recommend management strategies guided by inappropriate genetic inferences. Taken in isolation, this result suggests that students viewed HLADQ2 more deterministically than they should have, downplaying other possible causal agents. Such a response, predicted by two genetic essentialist biases – determinism and specific aetiology – are in line with Dar-Nimrod and Heine’s (2011) genetic essentialist framework. It also extends the literature on genetic essentialist biases from the focus lay populations (Dar-Nimrod et al., 2021) to medical professionals performing clinical reasoning. As the initial evidence for genetic essentialist biases playing a role in medical decision making, it raises concerns about potential serious diagnostic and management errors. Instances of cognitive bias leading to diagnostic error are not confined to the deterministic misinterpretation of genetic information. For instance, the misinterpretation of ANA positivity in the diagnosis of SLE is well recognised, since ANA positivity can be considered necessary (as ANA negative SLE is extremely rare) but insufficient for the diagnosis (Lesuis et al., 2017; Rajendran et al., 2021).

However, contrary to expectations, there was no significant difference between the two groups in their recommendations for follow up investigation. The majority of students across both groups responded (incorrectly) in-line with an investigation of coeliac disease, including those who received no genetic information in the patient vignette. This result may indicate a primary limitation to the testing question and/or the education the students received in reference to these kinds of clinical presentations. The directionality of the findings was in line with the hypothesis, so it may have been that insufficient statistical power obscured actual empirical support for the prediction.

These results are consistent with recent studies which suggest that medical students possess insufficient knowledge of genetics, especially around clinically orientated concepts such as genetic testing and genetic counselling (Alotaibi & Cordero, 2021). As such, greater integration of genetics into the clinical years of medical school curricula seems warranted, in line with a recent topic review from Wolyniak and colleagues, which concluded that there is a need for a development of scientific critical thinking skills, that allow students to apply foundational genetic knowledge and ethical principles to patient encounters (Wolyniak et al., 2015).

However, our qualitative analysis reveals a potential alternative account. The superordinate themes demonstrate that students have a raw, yet nuanced understanding of gene-action in the medical context. They acknowledge the non-deterministic influence of genes such as HLADQ2, and the importance of other causal forces for disease aetiology. The supportive quotes for superordinate theme 1, ‘the technique of passing the exam’ outline how students stated that the reason for their incorrect answers was not due to their lack of knowledge or reflective of their future clinical practice. Instead they indicated that they felt they needed to answer in line with the (perceived) examiners’ expectations, rather than the way they would act in clinical practice. The supportive quotes for superordinate theme 2 ‘being a holistic clinician’ outline how students expressed frustration with MCQs being insufficient to demonstrate their understanding of a topic in the clinical environment, which is a common criticism of this assessment modality (Anderson, 1979).

While the students demonstrated an awareness of the common non-deterministic gene-phenotype relationships in the qualitative analysis (superordinate theme 2 – my genetic understanding) this does not exclude the possibility that genetic essentialist biases played a role in decision making in Phase 1 of the study. While participants explicitly reasoned that the reasons for erroneous exam performance was because of conscious performative aspects of exam taking, it is possible that these post-hoc justifications do not accurately reflect the reasoning process that occurred at that time. Those deeper reflections may also arise once the students were informed about the true purpose of the study, and by extension, to their demonstration of a cognitive bias.

Another interesting finding from the qualitative analysis is that students perceive patients presenting with genetic information to be biased and fallible in their own understanding of that information. More research is required to understand how this perception might impact on their own clinical decision making. While diagnostic suggestions can bias medical decision making (Durning et al., 2012), it is not known whether suggestions that are perceived as biased, such as a patient’s genetic understanding, have the same effect.

Both perceived expectations (of assessors and patients) reflect a different aspect of genetic essentialist biases – the perception that *others* are engaging with genetic information in a biased manner. It has long been recognised that the provision of truthful and accurate information is necessary for patients to achieve sufficient health literacy beyond the merely functional level (Wittink & Oosterhaven, 2018). Erroneous interpretations of either patient-directed and acquired or practitioner-initiated genetic information may lead to flawed and deterministic decision-making, and become a barrier to achieving a truly interactive critical appraisal of one’s personal characteristics – a new ‘structural barrier to health’ (Nutbeam, 2000). Hence, the current findings have significant implications for both medical education and medical practice and warrant further research.

## Conclusion

The present findings indicate that genetic essentialist biases may affect clinical decision making of senior medical students. While the findings may also arise as an artifact of a conscious exam-taking strategy, there are substantial arguments against this interpetation.

## Data Availability

All data produced in the present study are available upon reasonable request to the authors

## Appendix 1 Phase 1 Questions

### Stem 1

Mary Sullivan is a 25 years-old female who presents to your GP surgery complaining of a 3-month worsening history of tiredness, intermittent diarrhoea, and generally “not feeling 100%”. Mary is worried because she has had the symptoms for a while now and asks, “could it be cancer?”. She has recently moved here from Adelaide, and hands you a summary of her medical history from her previous GP. You notice that she has previously had some genetic testing and has the HLADQ2 gene.

Please answer the following questions on gastroenterology, and Mrs Sullivan’s clinical presentation?

### Stem 2

Mary Sullivan is a 25 years-old female who presents to your GP surgery complaining of a 3-month history of tiredness, intermittent diarrhoea, and generally “not feeling 100%”. Mary is worried because she has had the symptoms for a while now and asks, “could it be cancer?”. She has recently moved here from Adelaide, and hands you a summary of her medical history from her previous GP.

Please answer the following questions on gastroenterology, and Mrs Sullivan’s clinical presentation?

Q1 Which of the following is TRUE with regards to the enteric nervous system?

A. The neurotransmitter noradrenaline stimulates peristalsis
B. Substance P relaxes the circular muscle
C. The vagus nerve has parasympathetic nervous system activity from oesophagus to the descending colon
D. Enterochromaffin cells are basal granular cells which release 5-HT with mucosal stimuli
E. It is situated predominantly between the serosa and the longitudinal muscle layer

Q2 Regarding the normal composition of faeces, which of the following is TRUE?

A. 75% is water
B. 40% is solid material
C. 50% of the solid material is dead bacteria
D. 10% of the solid material is protein
E. 50% of the solid material is fat

Q3 Which of the following is correct with regards to lipid digestion and absorption?

A. Pancreatic lipase causes hydrolysis of triglycerides
B. Fat malabsorption is confirmed when faecal fat is >3% of ingested fats
C. Approximately 60% of bile salts that enter the gut are reabsorbed in the terminal ileum
D. Micelles present triglycerides to the luminal membrane for absorption
E. Fat digestion fails when pancreatic function falls to < 20%

Q4 Regarding absorption in the gastrointestinal tract, which statement is TRUE?

A. Ethanol is absorbed after reaching the small intestine
B. Iron is absorbed in the proximal bowel as Fe^3+^
C. Sodium and glucose co-transporter on enterocytes is cAMP dependent
D. Carbohydrate absorption occurs predominantly in the distal ileum and proximal colon
E. Vitamin B12 is absorbed in the duodenum

Q5 Regarding pancreatic exocrine functions, which of the following is FALSE:

A. Trypsin inhibitor protects the pancreas from self-digestion
B. Zymogens are stored and released from pancreatic acinar cells
C. Trypsinogen is activated by enterokinase
D. Carboxypeptidases are involved in carbohydrate and protein digestion
E. Pancreatic insufficiency develops when 98% of the exocrine pancreas is lost

Q6 Regarding gastrointestinal tract motility, which of the following is TRUE?

A. Conscious muscular control of motility occurs at mouth, upper oesophageal sphincter, striated muscles of proximal oesophagus and anus
B. There is longer lag time in gastric emptying for liquids than solids to allow better absorption
C. Colonic High Amplitude Propagated Contractions occur during sleep
D. Phasic contractions allow the proximal stomach to act as storage
E. The main motility function of the small bowel is storage

Q7 Regarding this patient’s presenting history and symptoms, the next appropriate investigation would be?

A. Stool culture
B. Anti-tTG IgA antibody
C. Small bowel biopsy
D. Full Blood Count (FBC)
E. CT abdomen

Q8 Small intestine histological structures include all the following, EXCEPT?

A. Microvilli on intestinal side of enterocytes
B. Villi with a core of vasculature and lymphatics
C. Intraepithelial lymphocytes – predominantly T cells
D. Paneth cells with a defensive function
E. Crypts lined with columnar epithelium

Q9 Regarding this patient’s presenting history and symptoms, the most appropriate management would be?

A. Rehydration and recovery
B. Antibiotics
C. Referral to a dietician
D. Gluten free diet
E. Loperamide

Q10 In a patient presenting with symptoms that might suggest irritable bowel syndrome, which of the following would be considered a “red flag” that warrants further investigation?

A. A mixture of both diarrhoea and constipation
B. Symptoms developing < 45 years of age
C. Urgency and or feeling of incomplete evacuation
D. Nocturnal diarrhoea
E. History of anxiety/depression

